# Real-world evidence on levodopa dose escalation in patients with Parkinson’s disease treated with istradefylline

**DOI:** 10.1101/2022.06.02.22275921

**Authors:** Nobutaka Hattori, Daijiro Kabata, Shinji Asada, Tomoyuki Kanda, Takanobu Nomura, Ayumi Shintani, Akihisa Mori

## Abstract

**Objective:** Istradefylline, a selective adenosine A_2A_ receptor antagonist, is indicated in the US and Japan as adjunctive treatment to levodopa/decarboxylase inhibitors in adults with Parkinson’s disease (PD) experiencing OFF time. This study aimed to observe patterns of dose escalation of levodopa over time in patients initiated on istradefylline.

**Methods:** Using Japanese electronic health record data, interrupted time series analyses were used to compare levodopa daily dose (LDD, mg/day) gradients in patients before and after initiation of istradefylline. Data were analyzed by period relative to istradefylline initiation (Month 1): pre-istradefylline (Months -72 to 0), early istradefylline (Months 1 to 24), and late istradefylline (Months 25 to 72). Subgroup analyses included LDD before istradefylline initiation (<400, ≥400 to <600, ≥600 mg/day) and treatment with or without monoamine oxidase-B inhibitors (MAO-BIs), catechol-O-methyltransferase inhibitors (COMTIs), or dopamine agonists before istradefylline initiation.

**Results:** The analysis included 4026 patients; mean (SD) baseline LDD was 419.27 mg (174.19). Patients receiving ≥600 mg/day levodopa or not receiving MAO-BIs or COMTIs demonstrated a significant reduction in LDD increase gradient for pre-istradefylline vs late-phase istradefylline (≥600 mg/day levodopa, -6.259 mg/day each month, p<0.001; no MAO-BIs, -1.819 mg/day each month, p=0.004; no COMTIs, -1.412 mg/day each month, p=0.027).

**Conclusions:** This real-world analysis of Japanese prescription data indicated that slowing of LDD escalation was observed in patients initiated on istradefylline, particularly in those receiving ≥600 mg/day levodopa, suggesting istradefylline may slow progressive LDD increases. These findings suggest that initiating istradefylline before other levodopa-adjunctive therapies may mitigate LDD increases, potentially reducing occurrence or severity of levodopa-induced complications in long-term istradefylline treatment.

## Introduction

Although levodopa (LD) is the gold standard for Parkinson’s disease (PD) treatment [1], most patients receiving LD long-term eventually experience motor complications, including dyskinesia and wearing-off, causing declines in functional status and loss of LD benefit [2-4]. Disease duration and levodopa daily dose (LDD) have been identified as significant risk factors for development of motor complications [5,6], prompting use of the lowest possible LDD, with incremental increases made cautiously [1].

Istradefylline is a non-dopaminergic, selective adenosine A_2A_ receptor antagonist indicated in the US and Japan as adjunctive treatment to LD/decarboxylase inhibitors in adults with PD experiencing OFF episodes [7-9]. In a pooled analysis of eight 12- to 16-week randomized clinical trials, istradefylline (20 or 40 mg/day) resulted in a significant decrease in OFF time versus placebo, while ON time without troublesome dyskinesia significantly increased [10]. In a 1-year observational istradefylline post-marketing surveillance study, significant changes in LDD were not observed, prompting the question of whether adjunctive istradefylline could sufficiently treat symptoms, reducing the need for LDD increases [11].

Istradefylline’s adenosine A_2A_ receptor antagonism, a non-dopaminergic mechanism, has not historically been a standard PD treatment. In PD, balance between the basal ganglia’s direct and indirect pathways becomes disrupted when degeneration of nigrostriatal dopaminergic neurons causes loss of D_1_/D_2_ receptor-mediated control of both pathways [12]. Excitatory modulation via adenosine A_2A_ receptors expressed in the indirect pathway’s striatopallidal neurons is balanced by inhibitory dopamine D_2_ receptors in the same neuronal population [12]. In PD, attenuated D_2_ receptor-mediated inhibition and enhanced A_2A_ receptor-mediated excitation from increased receptor density result in excessive activation of the indirect output pathway, while reduced D_1_ receptor-mediated activation decreases direct pathway signaling, ultimately resulting in loss of motor control. Dopaminergic therapy *without* an A_2A_ receptor antagonist restores D_1_ stimulation, increasing striatonigral pathway activity, but it may not sufficiently restore the striatopallidal pathway’s D_2_ signaling [13]. When dopaminergic therapy is administered *with* an A_2A_ receptor antagonist, indirect pathway striatopallidal activity is reduced, leading to normalized movement control [13].

Although istradefylline may not be efficacious as monotherapy [14], once combined with even a low dose of LD, it may further improve motor symptoms without worsening motor complications, delaying the need for LDD increases over time. For example, in primate PD models, istradefylline significantly improved locomotor activity, reversed motor disability, and increased ON time in combination with a suboptimal LD dose [15,16]. In an exploratory clinical study, istradefylline and low-dose LD administered together to patients with PD potentiated LD response, with patients experiencing significantly less dyskinesia than those treated with higher-dose LD alone (p<0.05) [17].

We therefore consider istradefylline’s potential for reducing the need for LDD escalation via A_2A_ receptor antagonism and investigate its role in relieving motor complications or delaying their onset. Because the database used in this study did not provide information regarding wearing-off phenomena, a comparable control group of patients experiencing wearing-off but not receiving istradefylline treatment could not be established. Therefore, we used a self-controlled interrupted time series analysis (ITSA) design to explore patterns of LD dosage in patients with PD who were initiated on istradefylline therapy.

## Patients and methods

### Study design and data collection

This retrospective observational study captured anonymized hospital-based records, including prescription data, for more than 30 million patients from the diagnosis procedure combination (DPC) database (Medical Data Vision [MDV] Co., Ltd) in Japan between April 1, 2008, and April 30, 2019.

Eligible patients were diagnosed with PD (ICD-10 code G20 and disease codes 3320002, 8843950, 8843951, 8843952, 8843953, 8843954, or 8845602) and had a prescription for any LD formulation (including LD or LD/carbidopa, LD/benserazide, or LD/carbidopa/entacapone combinations) at least once during the observation period. Patients were not included if they were diagnosed with early-onset or familial PD (ICD-10 code G20, with disease codes 8841415, 8842319, 8843850, 8843851, 8843852, 8843853, 8843854, 8846156, 8846157, 8846158, or 8846159); secondary parkinsonism or other degenerative diseases of the basal ganglia (ICD-10 codes G21, G22, or G23); or only parkinsonism (ICD-10 codes G20 and disease codes 3320001 or 8830558 alone) during the record collection period or if their LD treatment initiation occurred before 40 years of age. The target population for analysis included patients who were prescribed istradefylline and continued for at least 29 days (defined as the period from the date of initial prescription to the date of the last prescription, plus one) from initial prescription.

Patient data were anonymized prior to incorporation into the MDV database, and the retrieved medical claims were therefore not linkable to individual patients. Additionally, this analysis did not require active enrollment, follow-up, or direct contact with patients. Therefore, institutional review board approval and written informed consent were not required for the conduct of this study.

### Outcomes

Study outcomes reported were changes in LDD (mg/day) and levodopa equivalent daily dose (LEDD, mg/day) before and after istradefylline initiation (**Fig 1**), assessed monthly. Dose adjustments in other dopaminergic medications, including dopamine agonists, MAO-B inhibitors, COMT inhibitors, and combinations, were broadly captured by LEDD data.

**Fig 1.**
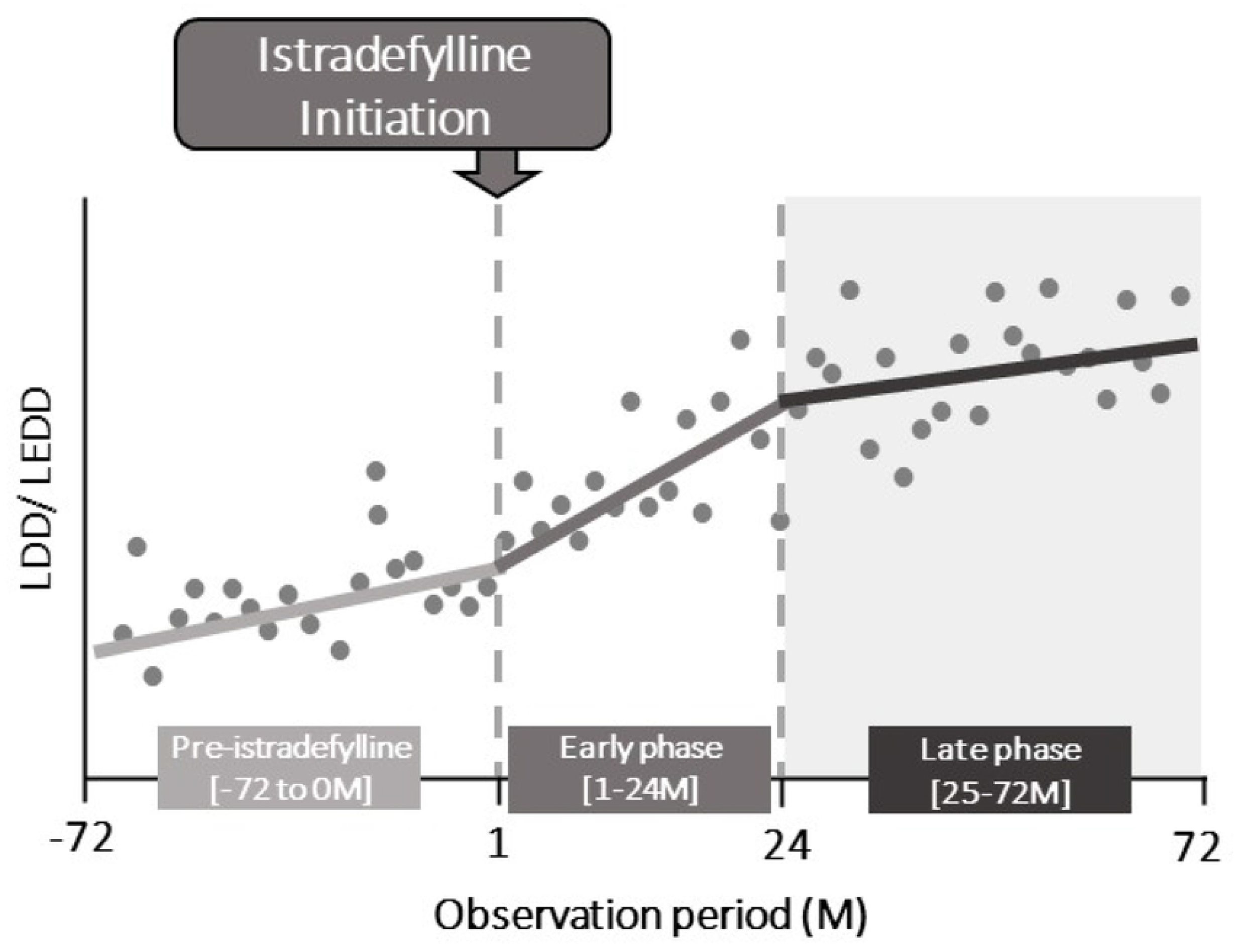
Study design for interrupted time series analysis study design. LEDD: levodopa equivalent daily dose; LDD: levodopa daily dose; M: month.

### Statistical analysis

We performed an ITSA using a segmented linear regression model to assess the effect of the initiation of istradefylline on the gradient changes in LDD and LEDD [18,19]. The gradient changes in LDD and LEDD can be interpreted as the average rate of the escalation in those values. The ITSA assessed each gradient change in LDD and LEDD within each observation period and examined whether the gradient changes varied between before and after istradefylline initiation. The observation periods were defined as follows: pre-istradefylline treatment period, Months -72 to 0; early istradefylline treatment period, Months 1 to 24 (early phase); and late istradefylline treatment period, Months 25 to 72 (late phase) (**Fig 1**). The early-phase and late-phase istradefylline treatment periods were arbitrarily determined based on the nonlinear relationship between the duration after Month 1 and the LD dosage change. In the segmented linear regression models for ITSA, we assumed that the LDD did not change immediately after the initiation of istradefylline treatment. Therefore, only the linear term of the gradient change was included, and terms relating to the level change intended for the acute effect of istradefylline treatment were excluded.

Subgroup analyses were conducted among subpopulations stratified by LDD (<400, ≥400 to <600, and ≥600mg/day) at istradefylline initiation, based on LDD at the time point immediately prior to istradefylline initiation, and treatment with a dopamine agonist, MAO-B inhibitor, or COMT inhibitor within three months prior to istradefylline initiation (defined as >2 prescriptions during months -2, -1, 0, and 1). The statistical significance level for hypothesis testing was two-sided 5%, and the confidence coefficient was 95%. Analyses were performed using R (https://www.r-project.org/).

## Results

### Patients

The analysis included 4026 patients (<400 mg/day LD, n=1733; ≥400 to <600 mg/day, n=1455; ≥600 mg/day, n=838). Baseline demographic characteristics were balanced with respect to gender and age (**Table 1**). Compared to patients treated with ≥600 mg/day LD, significantly fewer patients treated with lower doses of LD were receiving dopamine agonists and COMT inhibitors as concomitant medications (**Table 1**).

**Table 1.**
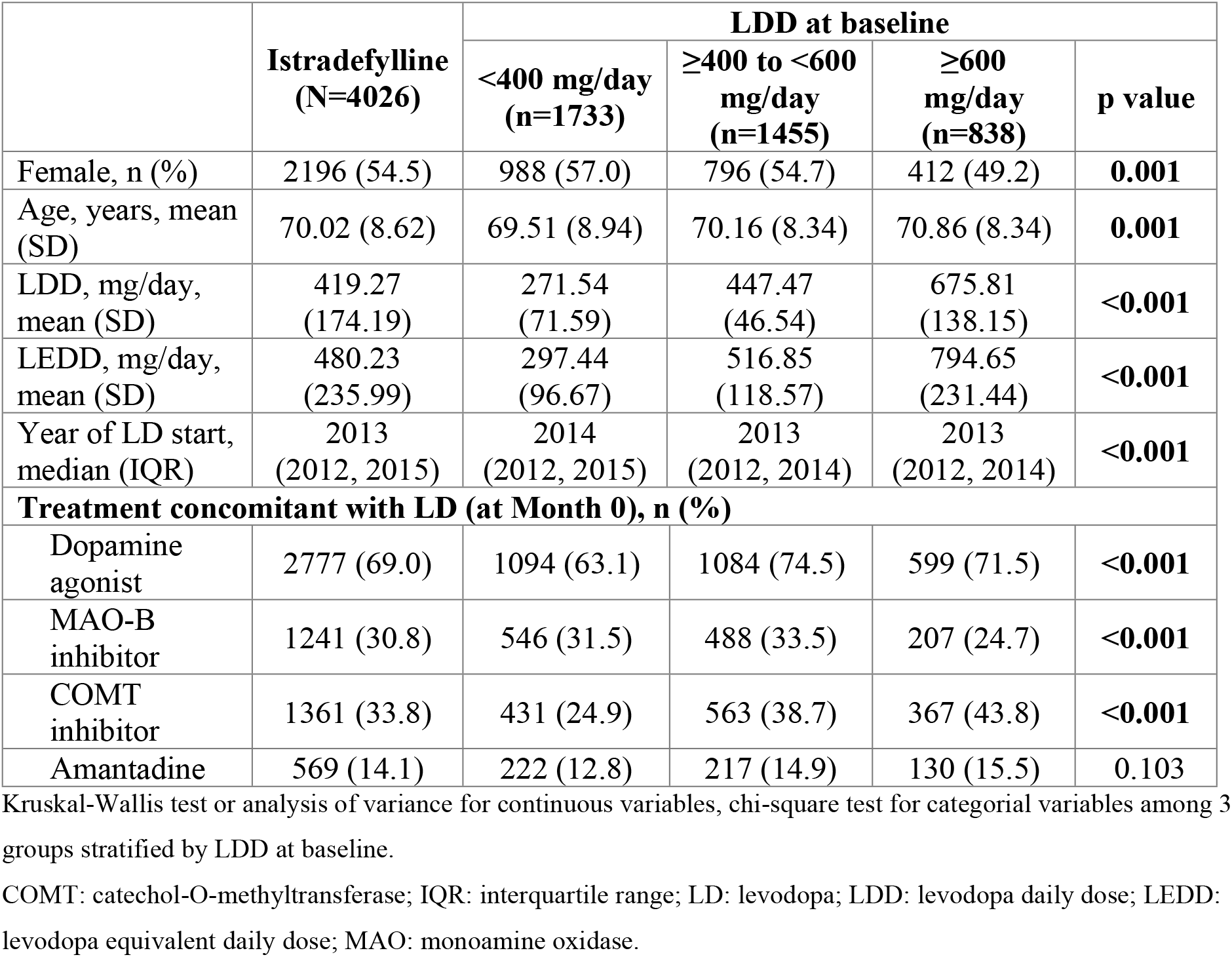
Baseline demographic and disease characteristics.

|  | **Istradefylline**<br>**(N=4026)** | **LDD at baseline** |  |  | **p value** |
| --- | --- | --- | --- | --- | --- |
|  |  | **<400 mg/day**<br>**(n=1733)** | **≥400 to <600**<br>**mg/day**<br>**(n=1455)** | **≥600**<br>**mg/day**<br>**(n=838)** |  |
| Female, n (%) | 2196 (54.5) | 988 (57.0) | 796 (54.7) | 412 (49.2) | **0.001** |
| Age, years, mean (SD) | 70.02 (8.62) | 69.51 (8.94) | 70.16 (8.34) | 70.86 (8.34) | **0.001** |
| LDD, mg/day, mean (SD) | 419.27 (174.19) | 271.54 (71.59) | 447.47 (46.54) | 675.81 (138.15) | **<0.001** |
| LEDD, mg/day, mean (SD) | 480.23 (235.99) | 297.44 (96.67) | 516.85 (118.57) | 794.65 (231.44) | **<0.001** |
| Year of LD start, median (IQR) | 2013 (2012, 2015) | 2014 (2012, 2015) | 2013 (2012, 2014) | 2013 (2012, 2014) | **<0.001** |
| **Treatment concomitant with LD (at Month 0), n (%)** |  |  |  |  |  |
| Dopamine agonist | 2777 (69.0) | 1094 (63.1) | 1084 (74.5) | 599 (71.5) | **<0.001** |
| MAO-B inhibitor | 1241 (30.8) | 546 (31.5) | 488 (33.5) | 207 (24.7) | **<0.001** |
| COMT inhibitor | 1361 (33.8) | 431 (24.9) | 563 (38.7) | 367 (43.8) | **<0.001** |
| Amantadine | 569 (14.1) | 222 (12.8) | 217 (14.9) | 130 (15.5) | 0.103 |

Kruskal-Wallis test or analysis of variance for continuous variables, chi-square test for categorical variables among 3 groups stratified by LDD at baseline.
COMT: catechol-O-methyltransferase; IQR: interquartile range; LD: levodopa; LDD: levodopa daily dose; LEDD: levodopa equivalent daily dose; MAO: monoamine oxidase.

### Impact of LDD at baseline on gradient of LDD and LEDD escalation

Among patients receiving ≥600 mg/day LD before istradefylline initiation, there were statistically significant reductions in the gradient of LDD escalation between the pre-istradefylline and early-phase treatment periods and between the pre-istradefylline and late-phase periods (**Fig 2**). These findings were consistent with the results for LEDD, which demonstrated significant gradient changes between the pre-istradefylline and early-phase periods of -3.814 (95% CI, -7.123, -0.505; p=0.024) and between the pre-istradefylline and late phase periods of - 8.455 (95% CI, -12.856, -4.053; p<0.001) (**Table 2**).

**Fig 2.**
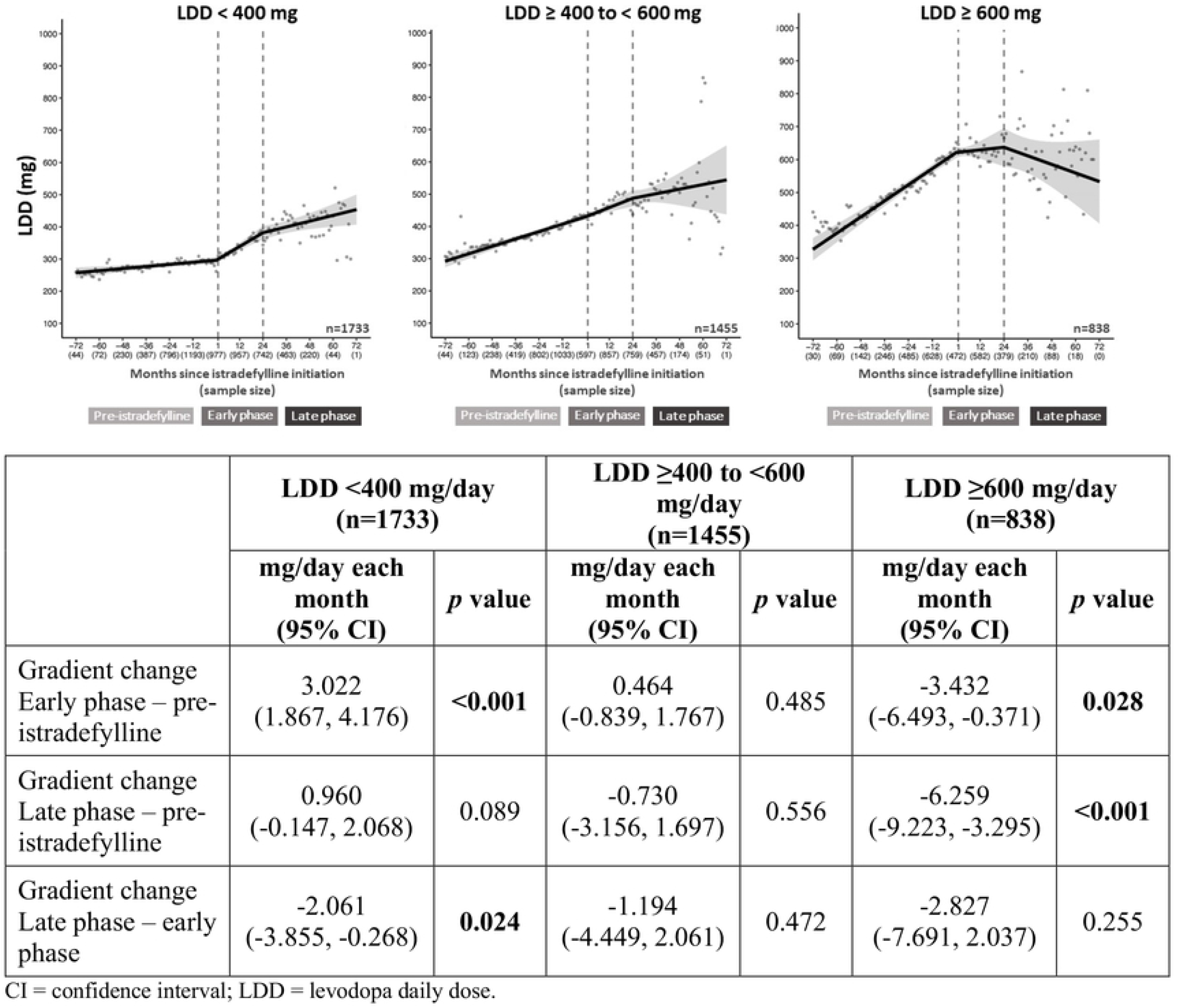
LDD escalation stratified by LDD at baseline. CI: confidence interval; LDD: levodopa daily dose.

**Table 2.** Stratification analysis for LEDD.

|  | Gradient changes in LEDD (per month) |  |  |
| --- | --- | --- | --- |
|  | Early phase - pre-istradefylline<br>mg/day each month<br>(95% CI) | Late phase – pre-istradefylline<br>mg/day each month<br>(95% CI) | Late phase – early phase<br>mg/day each month<br>(95% CI) |
| LDD at initiation<br><400 mg/day (n=1733) | **4.173**<br>**(2.571, 5.775)\*** | 1.475<br>(-0.139, 3.090) | **-2.698**<br>**(-5.281, -0.114)\*** |
| ≥400 to <600 mg/day<br>(n=1455) | 0.978<br>(-0.929, 2.886) | -0.170<br>(-3.505, 3.166) | -1.148<br>(-5.330, 3.034) |
| ≥600 mg/day (n=838) | **-3.814**<br>**(-7.123, -0.505)\*** | **-8.455**<br>**(-12.856, -4.053)\*** | -4.641<br>(-11.052, 1.770) |
| Treatment concomitant with LD (prior to Month 0) |  |  |  |
| With MAO-B inhibitor<br>(n=1150) | 0.995<br>(-1.543, 3.534) | 1.025<br>(-3.001, 5.501) | 0.030<br>(-5.206, 5.267) |
| Without MAO-B inhibitor<br>(n=2856) | 1.173<br>(-0.262, 2.607) | **-1.924**<br>**(-3.607, -0.240)\*** | **-3.096**<br>**(-5.594, -0.599)\*** |
| With COMT inhibitor<br>(n=1309) | -0.386<br>(-2.938, 2.165) | **-4.107**<br>**(-7.475, -0.739)\*** | -3.721<br>(-8.367, 0.925) |
| Without COMT inhibitor<br>(n=2697) | **2.476**<br>**(1.189, 3.762)\*** | -0.433<br>(-2.062, 1.197) | **-2.908**<br>**(-5.307, -0.509)\*** |
| With dopamine agonist<br>(n=2594) | 1.102<br>(-0.557, 2.761) | -0.760<br>(-3.102, 1.582) | -1.862<br>(-5.082, 1.357) |
| Without dopamine agonist<br>(n=1412) | 1.057<br>(-0.531, 2.645) | -2.156<br>(-4.665, 0.354) | -3.213<br>(-6.590, 0.165) |

\*p<0.05 by interrupted time series analysis.
COMT: catechol-O-methyltransferase; LD: levodopa; LDD: levodopa daily dose; LEDD: levodopa equivalent daily dose; MAO: monoamine oxidase.

In patients receiving <400 mg/day LD before initiating istradefylline, there was a significant increase in LDD gradient between the pre-istradefylline and early-phase periods, whereas a significant reduction in LDD gradient change occurred between the early-phase and late-phase periods (**Fig 2**). Similar results were found for the rate of LEDD escalation, with a gradient increase of 4.173 (95% CI, 2.571, 5.775; p<0.001) between the pre-istradefylline and early-phase periods and a decrease of -2.698 (−5.281, -0.114; p=0.041) between the early-phase and late-phase periods (**Table 2**).

No significant change in the LDD or LEDD gradient was observed between any treatment periods for patients treated with LD ≥400 and <600 mg/day (**Fig 2, Table 2**).

### Impact of concomitant dopaminergic medications (MAO-B inhibitors, COMT inhibitors, and dopamine agonists)

Among patients who did *not* receive MAO-B inhibitors prior to initiation of istradefylline, there was a significant decrease in LDD gradient change between the pre-istradefylline and late-phase periods and between the early-phase and late-phase periods (**Fig 3A**). For LEDD, there were significant decreases in escalation for patients who were *not* treated with MAO-B inhibitors prior to initiating istradefylline between the pre-istradefylline and late-phase periods and between the early-phase and late-phase periods (**Table 2**). In contrast, among patients who *received* MAO-B inhibitors, no significant difference in gradient change in LDD or LEDD was observed for any comparison.

**Fig 3.**
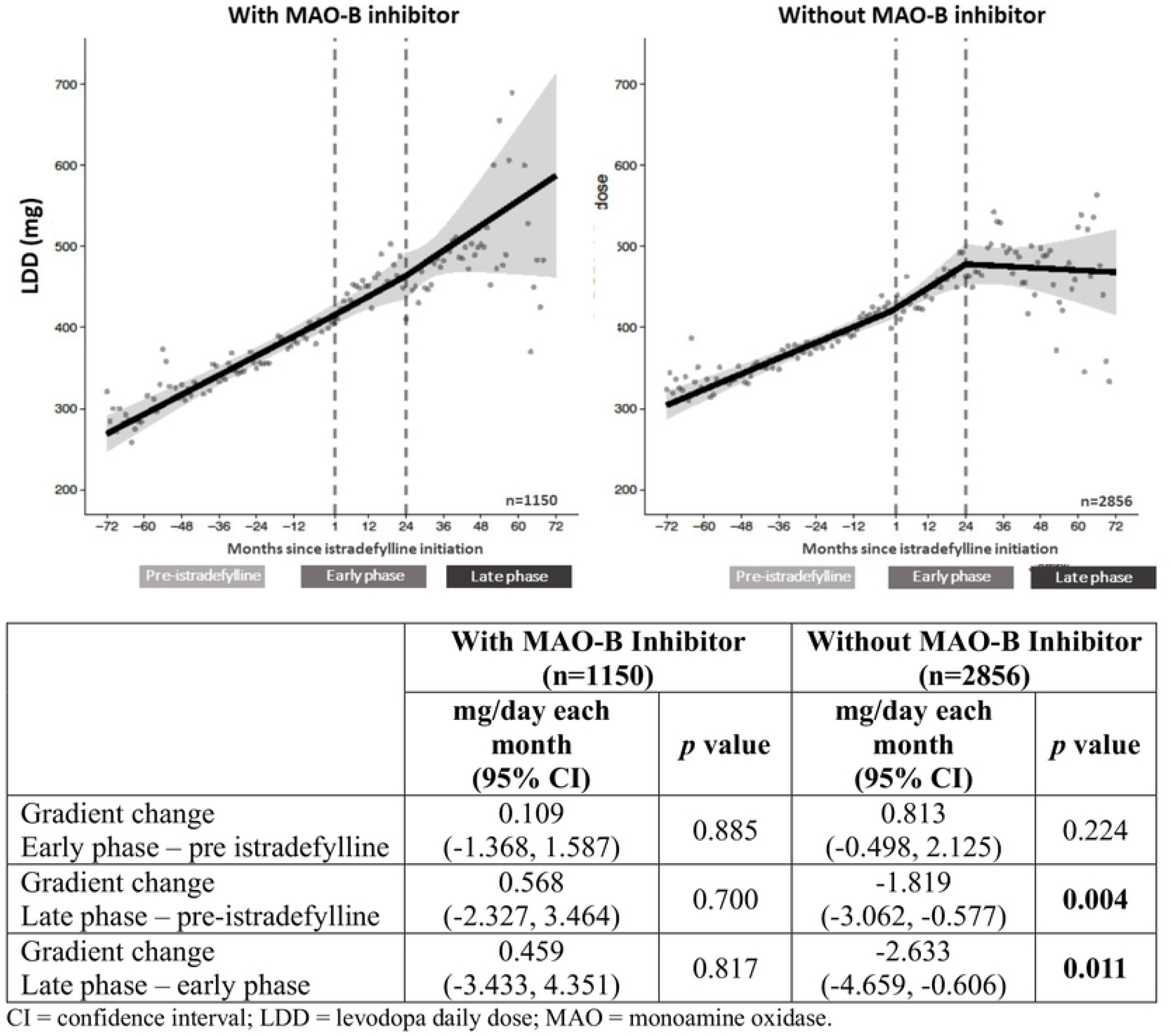
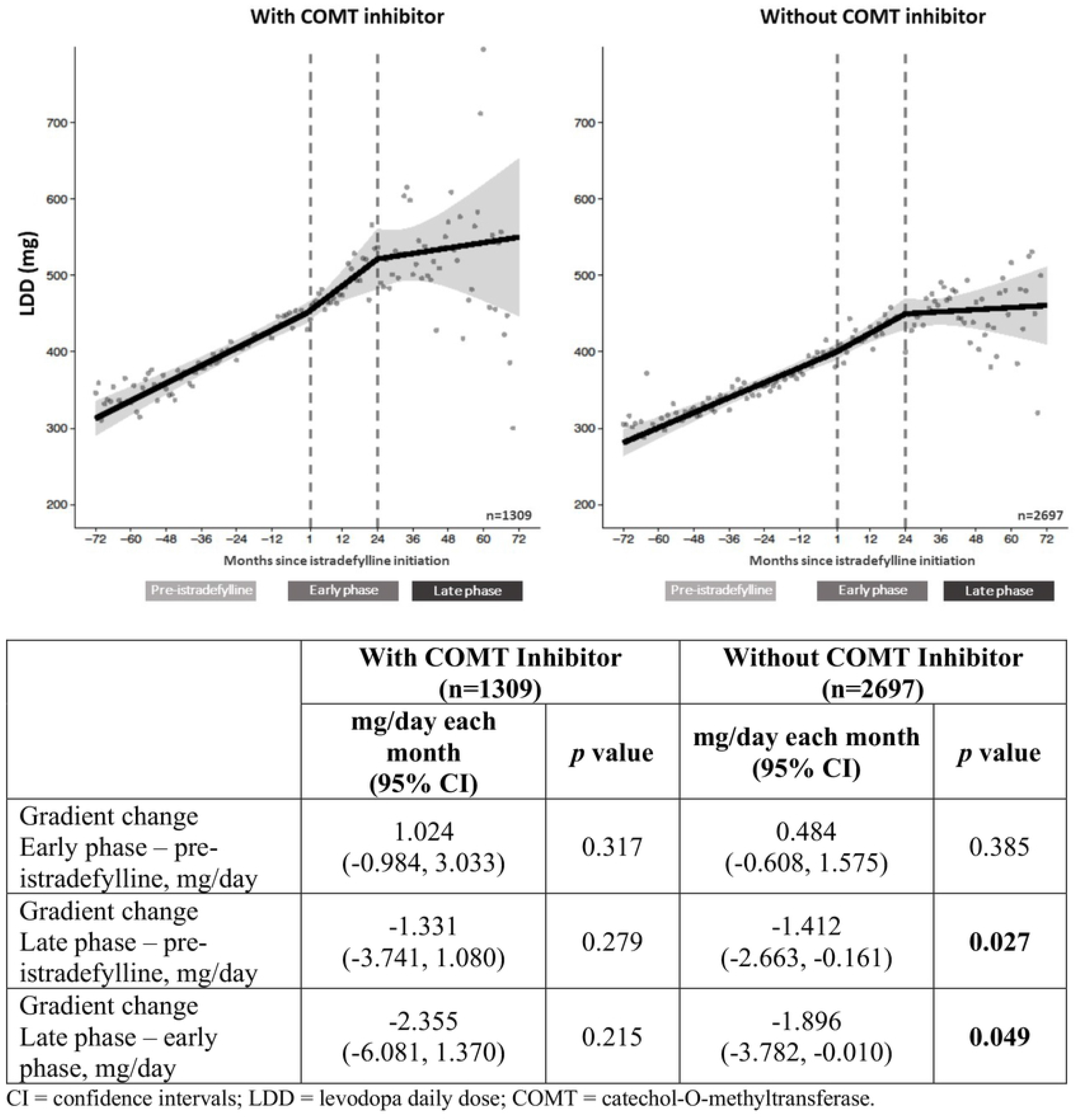
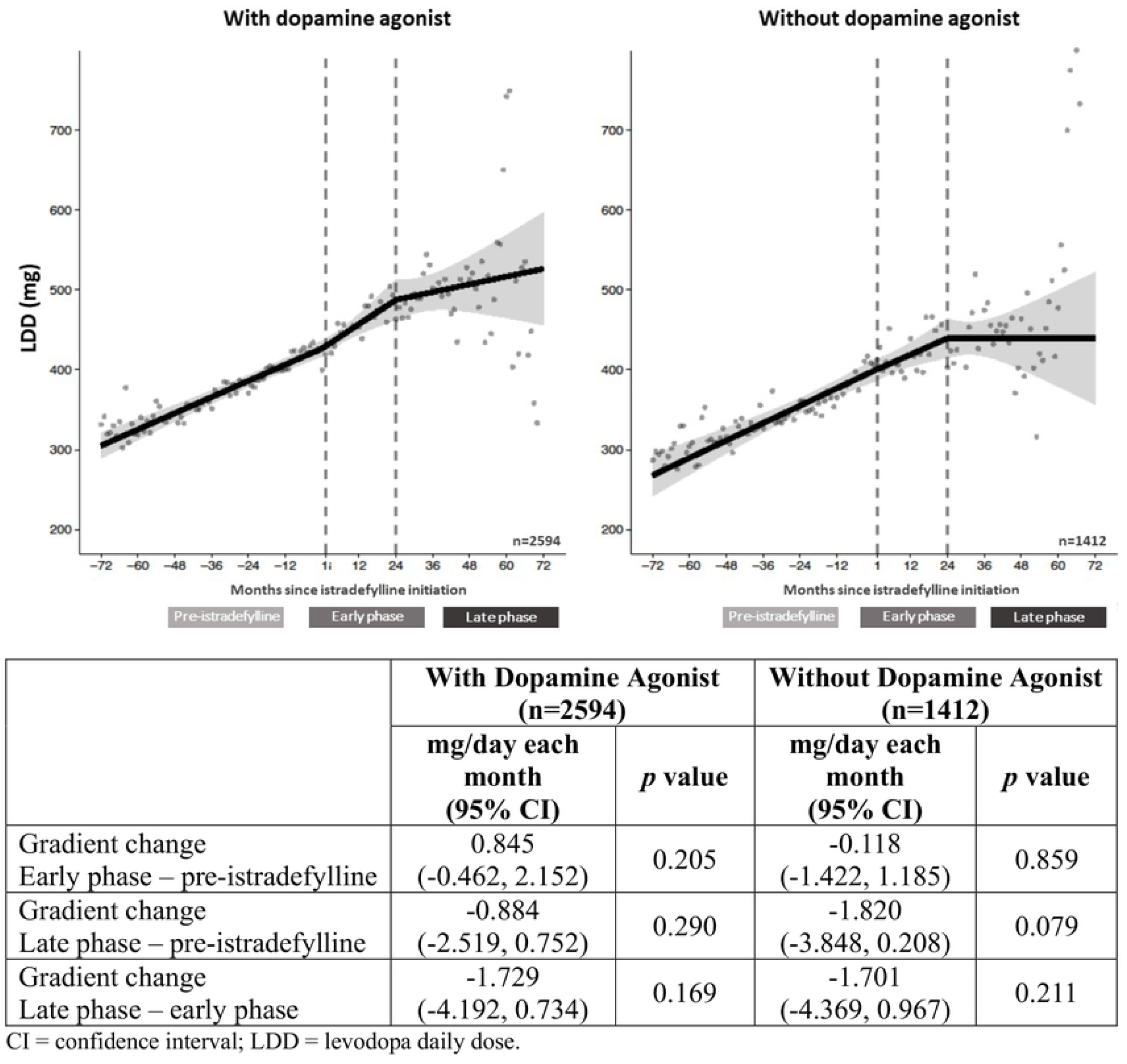
LDD escalation stratified by treatment with A) MAO-B inhibitors, B) COMT inhibitors, and C) dopamine agonists prior to istradefylline treatment. **A)** CI: confidence interval; LDD: levodopa daily dose; MAO: monoamine oxidase. **B)** CI: confidence interval; COMT: catechol-O-methyltransferase; LDD: levodopa daily dose. **C)** CI: confidence interval; LDD: levodopa daily dose.

For patients who did *not* receive COMT inhibitors prior to istradefylline initiation, statistically significant gradient decreases in LDD were observed between the pre-istradefylline and late-phase periods and the early-phase and late-phase periods (**Fig 3B**). By contrast, the rate of LEDD escalation significantly increased between the pre-istradefylline and early-phase periods and significantly decreased between the early-phase and late-phase periods (**Table 2**).

Patients who did *not* receive dopamine agonists prior to initiating istradefylline treatment demonstrated a similar but non-significant trend of negative change in LDD gradient, ie, suppression of dose escalation, between treatment periods as those who did not receive MAO-B or COMT inhibitors (**Fig 3C**).

## Discussion

In this real-world analysis of Japanese prescription data for patients with PD, an attenuation of the gradient of LDD increase could be observed after initiation of istradefylline therapy. This attenuation was found to persist for several years after istradefylline initiation. These observations suggest that the combination of istradefylline with LD may help maintain a therapeutic effect without necessitating further increases in LD dosage with disease progression. The negative shift in LDD gradient was most pronounced in patients receiving ≥600 mg/day LD at baseline, although this effect was also observed to a lesser extent in patients initially treated with <400 mg LD after two years of istradefylline treatment, as shown in **Fig 2**. A similar suppression of LEDD escalation was observed, indicating that the observed suppression of LDD escalation was not a result of a compensatory increase in LEDD. Statistically significant attenuations of LDD increase were observed in patients who had not received MAO-B inhibitors or COMT inhibitors before initiating istradefylline, with a similar, non-significant trend observed in patients who had not received dopamine agonists, as shown in **Fig 3**. Conversely, among patients who had received these adjunctive therapies prior to initiating istradefylline, no significant impact on suppression of LDD escalation over time was observed in all cases. These findings suggest that initiation of istradefylline treatment could more effectively slow progressive LDD increases in patients not receiving MAO-B inhibitors, COMT inhibitors, or dopamine agonists than in patients already treated with those adjunctive dopaminergic therapies. Among patients treated with LD at <600 mg/day (ie, the groups receiving <400 or ≥400 to <600 mg/day), no significant changes in LDD or LEDD gradients were observed. This finding warrants further investigation, including whether this observation may be attributable to Japanese physicians’ desire to avoid under-medication with LD to avoid increasing OFF episodes.

In this study, there was an apparent difference between early- and late-phase istradefylline treatment periods with regard to the gradient of LDD escalation. Among patients who received <600 mg/day LD at baseline, initiation of istradefylline did not have an appreciable impact on LDD escalation during the early-phase period. By contrast, among patients initially treated with ≥600 mg/day LD, a more pronounced suppression of LDD escalation was observed in the early-phase relative to the late-phase. Several twelve-week clinical trials of istradefylline that demonstrated significant reductions in OFF time and a trend towards improvement in motor symptoms had a median LD dose of ≥600 mg/day prescribed at baseline, whereas similar Japanese studies had a median LD dose of 400 mg/day [10]. However, LDD escalation may be required for further improvement, especially in motor symptoms, during the initial 24 months of istradefylline treatment. In this study, LDD in patients initially treated with <400 mg LD/day demonstrated a significant escalation for 24 months following istradefylline initiation, as shown in **Fig 2**, which may partly be driven by physicians seeking to achieve further improvement in motor symptoms.

In healthy individuals, excitatory modulation from adenosine A_2A_ receptors expressed in the striatopallidal neurons works in equilibrium with inhibitory dopamine D_2_ receptor-mediated regulation within the basal ganglia’s indirect pathway activity. In patients with PD, functional disruptions due to degeneration of dopaminergic inputs contribute to an imbalance between the direct and indirect pathways and loss of motor control [12]. This imbalance is thought to be amplified by an increase in A_2A_ receptor density with progression of PD, as a number of studies have shown an increase in A_2A_ receptor mRNA and adenosine binding sites in the striatum and globus pallidus external segment, particularly in patients with LD-induced motor complications compared with healthy subjects, as reviewed by Mori [13]. This pathophysiological change may contribute to the need for increases in dopaminergic treatments such as LD over the course of disease progression of PD. Although there is no study to date addressing whether A_2A_ receptor antagonists can induce a change in A_2A_ receptor expression patterns in the brain, it has been reported that expression of A_2A_ receptors in peripheral blood lymphocytes, which is increased in patients with PD (naïve to istradefylline treatment) [20], can be restored to levels comparable to those in healthy controls after 12 months of istradefylline treatment [21]. Furthermore, *ADORA2A*, the A_2A_ receptor gene, is subject to post-transcriptional silencing in individuals without PD [22,23]. Demethylation (activation) of *ADORA2A*, which has been observed in PD and may contribute to the observed increase in A_2A_ density, was reversed after 12 months of istradefylline treatment [21]. Although how or whether A_2A_ receptor dynamics in peripheral lymphocytes can be correlated with brain A_2A_ receptors remains to be investigated, based on these findings, it could be hypothesized that 1) over time, upregulation of A_2A_ receptor expression may contribute to symptomatic progression of PD, resulting from promoting imbalance of basal ganglia, and 2) the need to maintain a clinically effective level of dopamine stimulation by, for example, progressively increasing LDD, could be relieved by addition of istradefylline to selectively inhibit A_2A_ receptor function, thereby relieving motor dysfunction. On this basis, the istradefylline-induced epigenetic suppression of A_2A_ receptor upregulation may have interesting implications for the fundamental goals of LD therapy, although many areas remain to be investigated, including PET imaging of dopamine transporters and A_2A_ receptors in patients with PD receiving long-term istradefylline treatment [24-30]. If increased A_2A_ receptor expression in the indirect pathway contributes to the need for progressive increases in LDD, A_2A_ receptor epigenesis [21] data and the present real-world evidence (RWE) findings may explain the mechanism underpinning this correlation.

The real-world data used in this analysis were derived from a large database, with multiple years of data on LD and istradefylline treatment in patients with PD. Although the dataset was powerful for a study of this type, a limitation of the analysis is that the available data were observational, uncontrolled, and retrospective in nature and were confined to what was recorded in the database. Since no information was provided about patients’ clinical symptoms or disease severity or the presence or absence of motor complications (eg, wearing-off, dyskinesia), the clinical and prescribing criteria for LD dose adjustment are not known, and the database did not include efficacy or safety data. Therefore, the present analysis cannot directly determine how the attenuation of LDD increases in istradefylline-treated patients equates to a change in motor symptoms/complications or tolerability. Because of the lack of a controlled comparator or a propensity score matching analysis, which was not possible due to the limitations of the database, the patterns of LDD escalation over time in patients who did not initiate istradefylline treatment could not be determined. Furthermore, this analysis targeted a population who were prescribed and continued istradefylline for at least 29 continuous days; while it is likely that nearly all patients continued istradefylline for a significant period of time beyond this initial 29 days or even for the duration of the analysis, the mean treatment duration of istradefylline-treated patients from this database analysis could not be determined. A separate one-year observational study indicated that a similar population of Japanese patients with PD remained on istradefylline for a mean (SD) of 283.8 (130.7) days [11], and it can reasonably be assumed that the patients in this study would have had a similar pattern of istradefylline treatment. However, the absence of definitive data on duration of istradefylline treatment in the present analysis could limit the interpretation of the strength of any potential causal association between istradefylline treatment duration and change in pattern of LDD gradient change.

The results of this real-world analysis suggest that treatment with istradefylline may reduce the need for increases in LD dosage in PD patients over a period of several years. Further investigations concerning the mechanism underlying any potential effect of istradefylline on the rate of increase in LDD as well as the clinical consequences of long-term adjunctive therapy with LD and istradefylline, especially in terms of motor complications, are needed. Nevertheless, the present analysis provides new insights concerning long-term (ie, over 10 years) patterns of LDD before and after initiating istradefylline treatment. Future studies should evaluate the effect of initiation of istradefylline on symptoms associated with change/escalation of LDD treatment, such as the occurrence or severity of LD-induced motor complications. A study designed to evaluate whether adding istradefylline treatment delays the onset of dyskinesia in patients with PD receiving LD and experiencing wearing-off symptoms (Onset of Dyskinesia and Safety/Efficacy of Istradefylline [ODYSSEI]) is ongoing [31].

## Data Availability

The data used for the analyses in this manuscript are available on request from the corresponding author.

## Acknowledgments

Medical writing assistance was provided by Meghan Sullivan, PhD, and Michelle Jones, PhD, MWC, of MedVal Scientific Information Services, LLC, and was funded by Kyowa Kirin, Inc. (Princeton, NJ, USA).

## Author contributions

Conceptualization: Tomoyuki Kanda, Akihisa Mori

Data curation: Daijiro Kabata, Shinji Asada, Takanobu Nomura, Ayumi Shintani

Formal analysis: Daijiro Kabata

Funding acquisition: Tomoyuki Kanda, Akihisa Mori

Investigation: Nobutaka Hattori, Daijiro Kabata, Shinji Asada, Tomoyuki Kanda,

Takanobu Nomura, Ayumi Shintani, Akihisa Mori

Methodology: Tomoyuki Kanda, Akihisa Mori

Project administration: Tomoyuki Kanda

Resources: Tomoyuki Kanda

Software: Daijiro Kabata

Supervision: Nobutaka Hattori, Akihisa Mori

Validation: Daijiro Kabata, Shinji Asada, Takanobu Nomura, Ayumi Shintani

Visualization: Daijiro Kabata, Takanobu Nomura, Shinji Asada

Writing – original draft preparation: Akihisa Mori, Takanobu Nomura, Shinji Asada, Tomoyuki Kanda

Writing – review & editing: Nobutaka Hattori, Daijiro Kabata, Shinji Asada, Tomoyuki Kanda, Takanobu Nomura, Ayumi Shintani, Akihisa Mori

## Competing interests

**NH:** Research funding from AbbVie, Asahi Kasei Medical, Astellas, Boston Scientific, Daiichi Sankyo, Dai-Nippon Sumitomo, Eisai, FP Pharmaceutical, Kyowa Kirin, Medtronic, Mitsubishi Tanabe, MiZ, Nihon Medi-Physics, Nihon Pharmaceutical, Nippon Boehringer Ingelheim, OHARA, Ono Pharmaceutical, Otsuka, Takeda; consulting for Dai-Nippon Sumitomo; advisory boards for Dai-Nippon Sumitomo, Kyowa Kirin, Otsuka, Sanofi, Takeda; subcontracting (trial cases) for Biogen Japan, Hisamitsu, Meiji Seika; contract research for Biogen Japan, Mitsubishi Tanabe; honoraria from AbbVie, Alexion, Boston Scientific, Daiichi Sankyo, Dai-Nippon Sumitomo, FP Pharmaceutical, Kyowa Kirin, Lundbeck Japan, Medtronic, MSD K.K., Mylan, Nippon Boehringer Ingelheim, Novartis, Otsuka, Pfizer Japan, Takeda.

**DK:** Honoraria for statistical analysis, consulting, and lecturing from Kyowa Kirin Co., Ltd.

**SA, TK, TN & AM:** Employment with Kyowa Kirin Co., Ltd.

**AS:** Honoraria for statistical analysis, consulting, and lecturing from Kyowa Kirin Co., Ltd.

